## Appendix for "Syncope and subsequent traffic crash: A responsibility analysis"

**Supplemental appendix**

John A Staples, MD MPH [1, 2]

Shannon Erdelyi, MSc [3]

Ketki Merchant, MSc MBBS [1]

Candace Yip, BSc [1]

Mayesha Khan, MA [1]

Donald A Redelmeier, MD [4,5]

Herbert Chan, PhD [3]

Jeffrey R Brubacher, MD [2, 3]

[1] Department of Medicine, University of British Columbia, Vancouver, Canada
[2] Centre for Clinical Epidemiology & Evaluation, Vancouver, Canada
[3] Department of Emergency Medicine, University of British Columbia, Vancouver, Canada
[4] Sunnybrook Research Institute, Toronto, Canada
[5] Department of Medicine, University of Toronto, Toronto, Canada

Correspondence: John A Staples, MD MPH
VGH Research Pavilion, Room 276
828 West 10th Avenue, Vancouver BC V5Z1M9 Canada


**Syncope and subsequent traffic crash: A responsibility analysis**

**Supplemental Appendix**

**Table of contents**


| **Description** | **Page** |
|---|---|
| Item S1: Responsibility analysis accounts for changes in road exposure | 1 |
| Item S2: Data sources | 2 |
| Item S3: ICD diagnostic codes used to define baseline comorbidities | 3 |
| Item S4: Variable definitions | 5 |
| Item S5: Responsibility scores | 7 |
| Item S6: Crash characteristics among responsible and non-responsible drivers | 8 |
| Item S7: Forest plot of subgroup analyses | 12 |
| References | 13 |

### Item S1: Responsibility analysis accounts for changes in road exposure

| Variable | Scenario 1: Responsibility analysis of individuals with disease (~1% of population)<br><br>Road exposure identical among exposed and controls | Scenario 2: Responsibility analysis of individuals with disease (~1% of population)<br><br>Road exposure half as much among exposed relative to controls |
|---|---|---|
| **INPUT VALUES** | | |
| **Number of individuals** | | |
| Exposed | 1,000 | 1,000 |
| Control | 99,000 | 99,000 |
| **Crash risk (per 100,000 km traveled)** | | |
| Exposed | 0.20 | 0.20 |
| Control | 0.10 | 0.10 |
| **Travel distance per year (units of 100,000 km)** | | |
| Exposed | 1 | **0.50** |
| Control | 1 | 1 |
| **Proportion of crashes for which drivers are responsible** | | |
| Exposed | 0.75 | 0.75 |
| Control | 0.50 | 0.50 |
| **COMPUTED VALUES** | | |
| **Number of responsible crashes** | | |
| Exposed (C) | 150 | 75 |
| Control (D) | 4,950 | 4,950 |
| **Number of non-responsible crashes** | | |
| Exposed (E) | 50 | 25 |
| Control (F) | 4,950 | 4,950 |
| **Odds ratio** | 3 | 3 |

**Legend:** Table using hypothetical data to illustrate that responsibility analyses inherently account for road exposure because all crash-involved drivers were driving at the time of the crash. Grey cell highlights the change in inputs as the reader moves from left to right. Odds ratio = (C/D)÷(E/F). Scenario 1 depicts a responsibility analysis of all crashes in a population of 100,000 individuals, 1% of whom have a disease ('exposed'), with identical road exposure between exposed and control individuals. Scenario 2 depicts a responsibility analysis of all crashes among exposed and controls, but with a yearly travel distance among exposed that is half that of controls. The odds ratios produced in Scenarios 1 and 2 are identical and reflect only the relative proportion of crashes for which each group is responsible; they are unaffected by changes in cohort size and road exposure provided the probability of responsibility is not influenced by road exposure.

**Item S2: Data sources**

- **Laboratory data**: VCH Patient Care Information System [creator] (2018): Serum concentrations of hemoglobin, hematocrit, and troponin I. Vancouver Coastal Health [publisher]. Data Extract. VCH (2018).
- **Electrocardiogram (ECG) data**: VCH Regional MUSE™ Cardiology Information System v9 (General Electric, Boston, Massachusetts, USA) [creator] (2018): Numerical ECG data and physician ECG interpretation. Vancouver Coastal Health [publisher]. Data Extract. VCH (2018). Included all ECGs from the index ED visit and from baseline ECGs, defined as the two most recent ECGs in the VCH Regional MUSE™ system.
- **Consolidation File**: British Columbia Ministry of Health [creator] (2019): Consolidation File (MSP Registration & Premium Billing). V2. Population Data BC [publisher]. Data Extract. MOH (2018).
- **Medical Services Plan**: British Columbia Ministry of Health [creator] (2018): Medical Services Plan (MSP) Payment Information File. Population Data BC [publisher]. Data Extract. MOH (2018).
- **National Ambulatory Care Reporting System**: Canadian Institute for Health Information [creator] (2018): National Ambulatory Care Reporting System (NACRS). V2. Population Data BC [publisher]. Data Extract. MOH (2018).
- **Discharge Abstract Database**: Canadian Institute for Health Information [creator] (2019): Discharge Abstract Database (Hospital Separations). Population Data BC [publisher]. Data Extract. MOH (2018).
- **PharmaNet**: British Columbia Ministry of Health [creator] (2019): PharmaNet. V2. Population Data BC [publisher]. Data Extract. Data Stewardship Committee (2018).
- **Income Band**: Statistics Canada [creator]: Statistics Canada Income Band Data. Catalogue Number: 13C0016. V2. Population Data BC [publisher]. Data Extract. Population Data BC (2018).
- **Driver data** (Driver license, BC Traffic Accident System, ICBC Claims File): Insurance Corporation of British Columbia [creator] (2019): Driver Experience, Contraventions, and Exam tables and the Traffic Accident System. Insurance Corporation of British Columbia [publisher]. Data Extract. ICBC (2018).

We linked health and driving data using a previously established probabilistic linkage between Personal Health Number and Driver License Number based on name, sex and birthdate, with linkage rates exceeding 95%.[1]

All inferences, opinions and conclusions drawn in this manuscript are those of the authors and do not reflect the opinions or policies of the Data Stewards.

### Item S3: ICD diagnostic codes used to define baseline comorbidities

| Condition | Codes |
|---|---|
| **Myocardial infarction** | ICD9: 410, 412; ICD10: I21, I22, I252 |
| **Congestive heart failure** | ICD9: 39891, 402, 404, 425, 428; ICD10: I43, I50, I099, I110, I130, I132, I255, I420, I425, I426, I427, I428, I429, P290 |
| **Peripheral vascular disease** | ICD9: 0930, 437, 440, 441, 443, 4471, 5571, 5579, V434; ICD10: I70, I71, I731, I738, I739, I771, I790, I792, K551, K558, K559, Z958, Z959 |
| **Cerebrovascular disease** | ICD9: 36234, 430-438; ICD10: G45, G46, I60-I69, H340 |
| **Dementia** | ICD9: 290, 2941, 3312; ICD10: F00-F03, G30, F051, G311 |
| **Chronic obstructive pulmonary disease** | ICD9: 4168, 4169, 490-496, 500-505, 5064, 5081, 5088; ICD10: J40-J47, J60-J67, I278, I279, J684, J701, J703 |
| **Rheumatic disease** | ICD9: 4465, 7100-7104, 7140-7142, 7148, 725; ICD10: M05, M32-M34, M06, M315, M351, M353, M360 |
| **Peptic ulcer disease** | ICD9: 531-534; ICD10: K25-K28 |
| **Mild liver disease** | ICD9: 07022, 07023, 07032, 07033, 07044, 07054, 0706, 0709, 570, 571, 5733, 5734, 5738, 5739, V427; ICD10: B18, K73, K74, K700-K703, K709, K717, K713-K715, K760, K762- K764, K768, K769, Z944 |
| **Diabetes without complications** | ICD9: 2500-2503, 2508, 2509; ICD10: E100, E101, E106, E108, E109, E110, E111, E116, E118, E119, E120, E121, E126, E128, E129, E130, E131, E136, E138, E139, E140, E141, E146, E148, E149 |
| **Diabetes with complications** | ICD9: 2504-2507; ICD10: E102- E105, E107, E112-E115, E117, E122-E125, E127, E132-E135, E137, E142-E145, E147 |
| **Paraplegia and hemiplegia** | ICD9: 3341, 342, 343, 3440-3446, 3449; ICD10: G81, G82, G041, G114, G801, G802, G830-G834, G839 |
| **Renal disease** | ICD9: 403, 404, 582, 5830, 5831, 5832, 5834, 5836, 5837, 585, 586, 5880, V420, V451, V56; ICD10: N18, N19, N052-N057, N250, I120, I131, N032-N037, Z490-Z492, Z940, Z992 |
| **Cancer** | ICD9: 140-165, 170-172, 174-176, 179-195, 200- 208, 2386; ICD10: C00-C26, C30-C34, C37-C41, C43, C45-C58, C60-C76, C81-C85, C88, C90-C97 |
| **Moderate or severe liver disease** | ICD9: 4560-4562, 5722-5724, 5728; ICD10: K704, K711, K721, K729, K765-K767, I850, I859, I864, I982 |
| **Metastatic carcinoma** | ICD9: 196-199; ICD10: C77-C80 |
| **HIV** | ICD9: 042; ICD10: B20-B24 |
| **Syncope** | ICD9: 7802; ICD10: R55 |
| **Atrial fibrillation and flutter** | ICD9: 427; ICD10: I48 |
| **Other arrhythmias** | ICD9: 426, 427, 74686, 7850; ICD10: I44-I47, I49, R00 |
| **Presence of AICD** | ICD9: 99604, V4502, V5332; ICD10: Z9501, Z9502, Z4501, Z4502 |
| **Seizure disorders** | ICD9: 345, 78033; ICD10: G40, R5680 |
| **Obstructive sleep apnea and other sleep disorders** | ICD9: 307, 327, 3270, 32711, 32712, 3272, 32720, 32721, 32723, 32724, 32726, 32727, 32729, 3273, 32730, 32731, 32732, 32733, 32734, 32735, 32736, 32737, 32739, 32742, 32743, 3275, 32752, 32753, 32759, 3278, 7805, 78050, 78051, 78053, 78055-78059, V694; ICD10: F51, G47 |

### Item S3: ICD diagnostic codes used to define baseline comorbidities (continued)

| Condition | Codes |
|---|---|
| **Traumatic brain injury** | ICD9: 310, 80009, 80019, 80029, 80039, 80049, 80059, 80069, 80079, 80089, 80099, 80109, 80119, 80129, 80139, 80149, 80159, 80169, 80179, 80189, 80199, 80309, 80319, 80329, 80339, 80349, 80359, 80369, 80379, 80389, 80399, 80409, 80419, 80429, 80439, 80449, 80459, 80469, 80479, 80489, 80499, 850, 8500, 85011, 85012, 8502-8505, 8509, 85109, 85119, 85129, 85139, 85149, 85159, 85169, 85179, 85189, 85199, 85209, 85219, 85229, 85239, 85249, 85259, 85309, 85319, 85409, 85419, V1552, V8001; ICD10: F072, S06 |
| **Psychiatric disorders** | ICD9: 295-301, 306-319; ICD10: F04-F09, F2-F9 |
| **Alcohol misuse** | ICD9: 291, 303, 305, 3575, 425, 5353, 53530, 53531, 571, 76071, 7903, 9773, 980, E860, V113; ICD10: F10, G312, G721, I426, K292, K70, K852, O354, R780, T51, X45, X65, Y15, Y90, Y91, Z502, Z714, Z721, Z8640 |
| **Other substance misuse** | ICD9: 292, 304, 305, 76073, 76075, 9650, 96501, 96502, 96509, 967, 9670, 9671, 9676, 9678, 9679, 9696, 96972, 9701, 97081, E8500, E8501, E8502, E851, E852, E8520, E8525, E8528, E8529, E8541, E9350, E9351, E9352, E937, E9370, E9371, E9376, E9378, E9379, E9396, E9401, E9501, E9502, E9801, E9802; ICD10: F11-F19 |
| **Chronic ischemic heart disease** | ICD9: 414; ICD10: I25 |
| **Hypertension** | ICD9: 36211, 401-405, 6420, 64200-64204, 6421, 64210-64214, 6422, 64220-64224, 6427, 64270-64274; ICD10: I10-I15, O10, O11 |
| **Pacemaker** | ICD9: V4501, V5331; ICD10: Z4500, Z9500, Z9502, Z4502 |
| **Unstable angina** | ICD10: I200 |
| **Cardiovascular disease (CVD)** | Codes from myocardial infarction, chronic ischemic heart disease, peripheral vascular disease, congestive heart failure, atrial fibrillation and flutter, other cardiac arrhythmia, cerebrovascular disease |

**Legend:** We considered comorbidities present if identified in diagnostic coding from ≥1 hospitalization or ≥2 physician visits in a 1-year lookback interval. ICD = The World Health Organization's International Statistical Classification of Diseases and Related Health Problems; ICD9 = ICD, 9th Revision, Clinical Modification (ICD-9-CM) codes; ICD10 = ICD, 10th Revision, Canada (ICD-10-CA) codes; HIV = human immunodeficiency virus; AICD = automated internal cardioverter-defibrillator, CVD = cardiovascular disease.

### Item S4: Variable definitions

| Variables | Definition |
|---|---|
| **Outcome** | |
| Crash responsibility | Crash responsibility was determined using objective police data and a validated responsibility scoring tool. Responsibility scores were categorized as 'responsible' (score ≤13), 'non-responsible' (score ≥16), or indeterminate (score 14 - 15) in the main analysis. |
| **Exposures** | |
| Syncope | An emergency department visit with a discharge diagnosis of 'syncope and collapse' that occurred in the exposure lookback interval. The main analysis used a 3-monoth exposure lookback interval. |
| Syncope 'definite or likely' | Among individuals with syncope as defined above, those in whom syncope was deemed by trained chart abstractors to be 'definite' or 'very likely' based on comprehensive review of medical records from the index emergency department visit. |
| **Potential confounders** | |
| **Demographic** | |
| Age group | Driver age at index crash date (16-35 years; 36-55 years*; ≥56 years) |
| Sex | Driver sex (male; female*) |
| Residential neighbourhood household income quintile | Driver residential neighbourhood income quintile generated by PopDataBC using census data and driver residential postal code (1 = lowest*, 5 = highest; ordinal) |
| **Driving history** | |
| License type | Type of driver license held at the time of index crash (full vs novice or learner*) |
| Years with automobile insurance in past 5y | Number of years with auto insurance in the 5 years before index crash (0 years*, >0 to ≤ 2.5 years, 2.5 to 5 years) |
| ≥1 contravention in the past 5y | Traffic violations include speeding, distracted driving, or impaired driving in the 5 years before index crash (yes/no). |
| ≥1 crash in the past 5y | Police-reported crash in the 5 years before index crash (yes/no). |
| **Crash characteristics** | |
| Crash season | Season of index crash: Winter (Dec- Feb); Spring (Mar- May); Summer (Jun-Aug); Fall (Sep-Nov)* |
| Substance impairment at time of crash | Breath or blood test positive for alcohol, or alcohol or drug impairment suspected by police officer (yes/no) |
| Annual percent responsible by crash year | Of all police-reported crashes in BC, the proportion of crashes deemed 'responsible' in the index crash year (continuous variable)[1] |
| **Health** | |
| CCI in the past year | Dichotomized variable for Charlson Comorbidity Index (CCI) ≥2 (yes/no); Comorbidities were deemed to be present if the associated diagnostic codes were found in ≥1 hospitalization or ≥2 clinic visits in the year before index crash |
| Hospitalization in the past year | Dichotomized variable for ≥1 hospitalizations in the year before index crash (yes/no). |
| Physician visits in the past year | Number of physician/clinic visits in the year before index crash; 0 visits*, ≤ 1 per month (1-12 visits); ≤ 1 per week (13-52 visits); > 1 per week (>52 visits) |
| History of cardiovascular disease in past year | ≥1 hospitalization or ≥2 clinic visits for cardiovascular disease in the year before index crash (yes/no) |
| History of diabetes in past year | ≥1 hospitalization or ≥2 clinic visits for diabetes in the year before index crash (yes/no) |
| History of alcohol misuse in past year | ≥1 hospitalization or ≥2 clinic visits for alcohol use disorder in the year before index crash (yes/no) |
| History of other substance misuse in past year | ≥1 hospitalization or ≥2 clinic visits for other substance use disorder in the year before index crash (yes/no) |

| | |
|---|---|
| Presence of AICD or pacemaker in past year | Presence of diagnostic codes for automated internal cardioverter-defibrillator (AICD) or pacemaker in hospital or clinic visit records in the year before index crash (yes/no) |
| Number of prescription medications prescribed within 60 days of crash | Number of distinct prescription medications likely consumed in the 60 days prior to index crash excluding those prescribed on the index crash date (0*, 1, ≥2 distinct medications). Overlap with 60-day lookback determined using dispensation date and days supplied. |
| Benzodiazepines prescribed within 60 days of crash | Prescription for benzodiazepines dispensed in the 60 days prior to index crash, excluding index crash date (yes/no). |
| Opioids prescribed within 60 days of crash | Prescription for opioids dispensed in the 60 days prior to index crash, excluding index crash date (yes/no). |

**Legend:** * indicates the referent category used in regression analyses.

**Item S5: Responsibility scores**

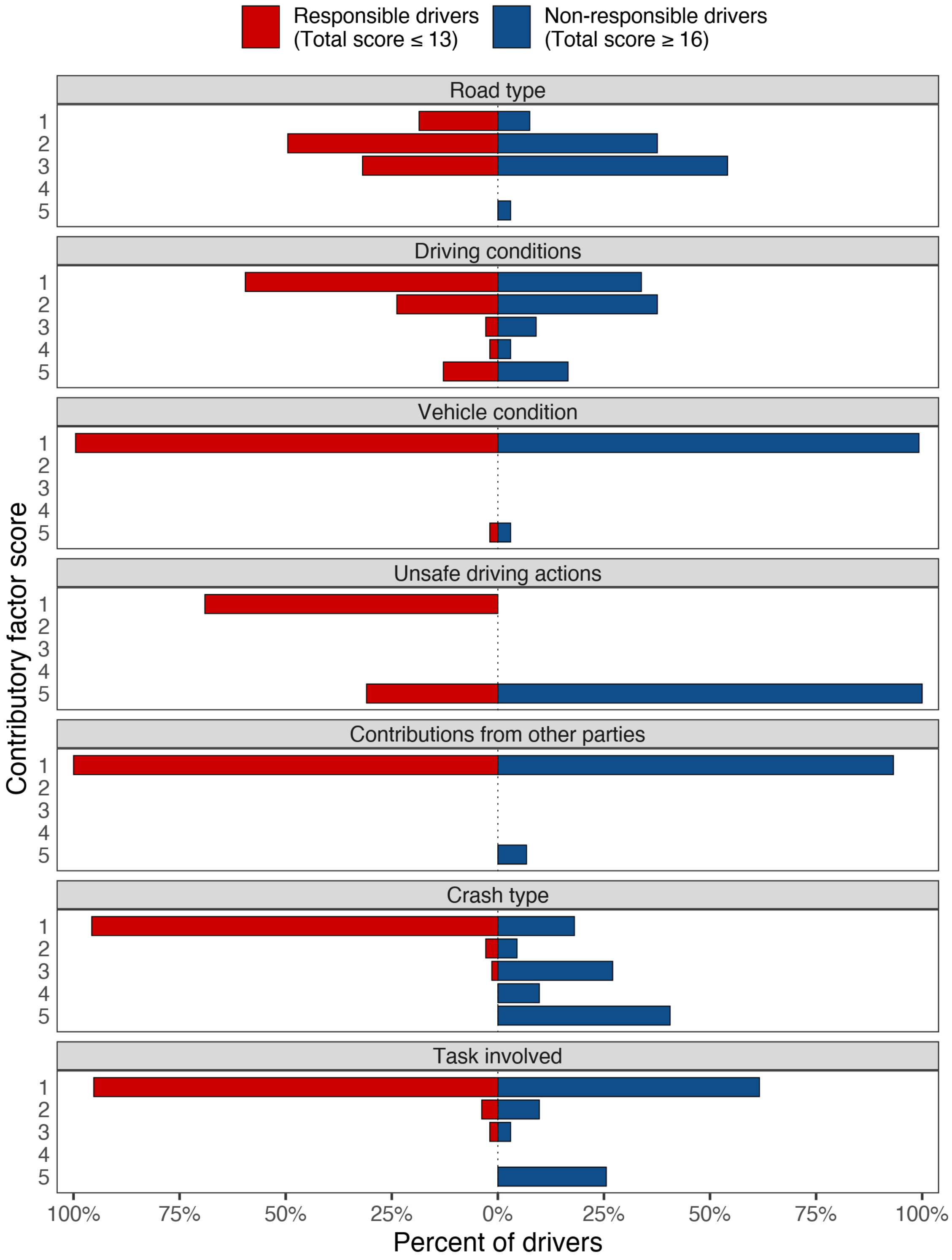


**Legend:** Mirrored bar chart comparing scores between responsible and non-responsible drivers for each of the seven components of the responsibility score.

### Item S6: Crash characteristics among responsible and non-responsible drivers

| Characteristic | Responsible n = 210 (100%) | Non-responsible n = 133 (100%) | Indeterminate n = 132 (100%) | p-value (responsible vs. non-responsible) |
|---|---|---|---|---|
| **Road type** | | | | |
| Traffic flow | | | | |
| One-way | 32 (15.2%) | 9 (6.8%) | 12 (9.1%) | 0.02 |
| Two-way | 171 (81.4%) | 123 (92.5%) | 116 (87.9%) | |
| Other/unknown | 7 (3.3%) | < 5 | < 5 | |
| Road class | | | | |
| Single lane | 6 (2.9%) | 6 (4.5%) | 7 (5.3%) | 0.02 |
| Single lane, ramp | | | | |
| Multi-lane | 159 (75.7%) | 106 (79.7%) | 106 (80.3%) | 0.02 |
| Multi-lane, ramp | 0 | 0 | < 5 | |
| Not applicable | 30 (14.3%) | 6 (4.5%) | < 5 | |
| Other/unknown | 15 (7.1%) | 15 (11.3%) | 14 (10.6%) | |
| Accident location | | | | |
| At intersection | 73 (34.8%) | 75 (56.4%) | 56 (42.4%) | <0.0001 |
| Between intersection | 74 (35.2%) | 45 (33.8%) | 48 (36.4%) | |
| Parking lot | 22 (10.5%) | < 5 | < 5 | |
| Other/unknown | 41 (19.5%) | 11 (8.3%) | 25 (18.9%) | |
| Roadside hazard or road design listed as contributory factor | 0 | < 5 | 0 | 0.82 |
| **Driving conditions** | | | | |
| Road condition | | | | |
| Dry | 137 (65.2%) | 52 (39.1%) | 97 (73.5%) | <0.0001 |
| Wet | 62 (29.5%) | 70 (52.6%) | 28 (21.2%) | |
| Snow/slush/ice/mud | 9 (4.3%) | 10 (7.5%) | < 5 | |
| Other/unknown | < 5 | < 5 | < 5 | |
| Road surface | | | | |
| Asphalt/concrete | 208 (99.0%) | 133 (100.0%) | 130 (98.5%) | 0.69 |
| Stone/gravel/earth/wood | < 5 | 0 | < 5 | |
| Other/unknown | 0 | 0 | < 5 | |
| Weather | | | | |
| Clear/cloudy | 161 (76.7%) | 76 (57.1%) | 110 (83.3%) | 0.003 |
| Rain/strong wind | 38 (18.1%) | 48 (36.1%) | 16 (12.1%) | |
| Fog/smoke/smog | < 5 | < 5 | 0 | |
| Snow/sleet/hail | 5 (2.4%) | 5 (3.8%) | < 5 | |
| Other/unknown | < 5 | < 5 | < 5 | |
| Lighting | | | | |
| Daylight | 145 (69.0%) | 84 (63.2%) | 99 (75.0%) | 0.49 |
| Dusk/dawn | 18 (8.6%) | 8 (6.0%) | 5 (3.8%) | |
| Dark with full illumination | 20 (9.5%) | 17 (12.8%) | 17 (12.9%) | |
| Dark with no/some illumination | 26 (12.4%) | 23 (17.3%) | 10 (7.6%) | |
| Other/unknown | < 5 | < 5 | < 5 | |
| Weather or visibility listed as contributory factor | 27 (12.9%) | 22 (16.5%) | < 5 | 0.43 |
| **Vehicle condition** | | | | |
| Vehicle condition not listed as contributory factor | 209 (99.5%) | 132 (99.2%) | 131 (99.2%) | 1.00 |
| **Unsafe driving actions** | | | | |
| Index driver driving safely and | 65 (31.0%) | 133 (100.0%) | 132 (100.0%) | <0.0001 |

| | | | | |
|---|---|---|---|---|
| obeying road laws | | | | |
| Index driver not driving safely or disobeying road laws | 145 (69.0%) | 0 | 0 | |
| **Contributions from other parties** | | | | |
| Yes | 0 | 9 (6.8%) | 0 | 0.001 |
| No / index driver driving unsafely | 210 (100.0%) | 124 (93.2%) | 132 (100.0%) | |
| **Crash type** | | | | |
| Number of vehicles involved | | | | |
| Single vehicle | 55 (26.2%) | 23 (17.3%) | 12 (9.1%) | 0.02 |
| Two vehicles | 116 (55.2%) | 70 (52.6%) | 93 (70.5%) | |
| Three or more vehicles | 39 (18.6%) | 40 (30.1%) | 27 (20.5%) | |
| Diagram description | | | | |
| Intersection - right angle | 27 (12.9%) | 34 (25.6%) | 19 (14.4%) | 0.004 |
| Head on | 22 (10.5%) | 8 (6.0%) | 18 (13.6%) | |
| Rear end | 53 (25.2%) | 49 (36.8%) | 14 (10.6%) | |
| Backing | 6 (2.9%) | < 5 | < 5 | |
| Turn | 23 (11.0%) | 11 (8.3%) | 30 (22.7%) | |
| Overtaking | 8 (3.8%) | < 5 | 13 (9.8%) | |
| Off road | 23 (11.0%) | 8 (6.0%) | 7 (5.3%) | |
| Other/unknown | 48 (22.9%) | 18 (13.5%) | 29 (22.0%) | |
| Damage location | | | | |
| Front | 117 (55.7%) | 37 (27.8%) | 48 (36.4%) | <0.0001 |
| Front/rear | 10 (4.8%) | 5 (3.8%) | 5 (3.8%) | |
| Front/side | 10 (4.8%) | 10 (7.5%) | 17 (12.9%) | |
| Rear | 10 (4.8%) | 37 (27.8%) | 17 (12.9%) | |
| Rear/side | < 5 | < 5 | < 5 | |
| Side | 5 (2.4%) | 0 | < 5 | |
| Tires/undercarriage/ windshield/roof | 13 (6.2%) | 10 (7.5%) | 10 (7.6%) | |
| Whole vehicle | 8 (3.8%) | 7 (5.3%) | 8 (6.1%) | |
| None | 10 (4.8%) | 8 (6.0%) | 7 (5.3%) | |
| Other/unknown | 117 (55.7%) | 37 (27.8%) | 48 (36.4%) | <0.0001 |
| Pre-collision action | | | | |
| Straight | 124 (59.0%) | 81 (60.9%) | 81 (61.4%) | <0.0001 |
| Backing | 7 (3.3%) | < 5 | < 5 | |
| Turning | 49 (23.3%) | 12 (9.0%) | 36 (27.3%) | |
| Changing lanes/merging | < 5 | < 5 | < 5 | |
| Loss of control | 6 (2.9%) | 0 | < 5 | |
| Stopped/parked | 0 | 26 (19.5%) | < 5 | |
| Other/unknown | 20 (9.5%) | 11 (8.3%) | 7 (5.3%) | |
| **Task involved** | | | | |
| Avoidance manoeuvre listed as contributory factor | < 5 | 8 (6.0%) | < 5 | 0.005 |
| Pre-collision action | | | | |
| Straight | 124 (59.0%) | 81 (60.9%) | 81 (61.4%) | <0.0001 |
| Backing | 7 (3.3%) | < 5 | < 5 | |
| Turning | 49 (23.3%) | 12 (9.0%) | 36 (27.3%) | |
| Changing lanes/merging | < 5 | < 5 | < 5 | |
| Loss of control | 6 (2.9%) | 0 | < 5 | |
| Stopped/parked | 0 | 26 (19.5%) | < 5 | |
| Other/unknown | 20 (9.5%) | 11 (8.3%) | 7 (5.3%) | |
| **Other crash characteristics** | | | | |
| Human condition listed as contributory factor | 114 (54.3%) | 22 (16.5%) | 44 (33.3%) | <0.0001 |
| Alcohol | 11 (5.2%) | < 5 | < 5 | |

| | | | | |
|---|---|---|---|---|
| Medications | < 5 | 0 | 0 | |
| Drugs | < 5 | 0 | < 5 | |
| Illness/fatigue | 22 (10.5%) | < 5 | 7 (5.3%) | |
| Distracted/inattentive | 78 (37.1%) | 16 (12.0%) | 31 (23.5%) | |
| Pre-Existing Physical Disability | < 5 | < 5 | < 5 | |
| Breath alcohol positive | | | | |
| Yes | < 5 | 0 | < 5 | 1.00 |
| No/not tested | 209 (99.5%) | 133 (100.0%) | 131 (99.2%) | |
| Accident severity | | | | |
| Casualty (fatal or injury) | 104 (49.5%) | 63 (47.4%) | 63 (47.7%) | 0.78 |
| Property damage | 106 (50.5%) | 70 (52.6%) | 69 (52.3%) | |
| Road location | | | | |
| Rural road | < 5 | 0 | < 5 | 0.43 |
| Provincial highway | 49 (23.3%) | 25 (18.8%) | 15 (11.4%) | |
| City street | 160 (76.2%) | 108 (81.2%) | 116 (87.9%) | |
| Speed zone | | | | |
| < 50 km/h | 14 (6.7%) | < 5 | < 5 | 0.18 |
| 50 km/h | 117 (55.7%) | 89 (66.9%) | 101 (76.5%) | |
| 60 - 70 km/h | 24 (11.4%) | 12 (9.0%) | 13 (9.8%) | |
| >= 80 km/h | 29 (13.8%) | 17 (12.8%) | 5 (3.8%) | |
| Other/unknown | 26 (12.4%) | 12 (9.0%) | 10 (7.6%) | |
| Road character | | | | |
| Straight - flat | 128 (61.0%) | 81 (60.9%) | 75 (56.8%) | 0.20 |
| Straight - graded | 46 (21.9%) | 26 (19.5%) | 34 (25.8%) | |
| Curved - flat | | | | |
| Curved - graded | | | | |
| Other/unknown | 9 (4.3%) | 14 (10.5%) | 7 (5.3%) | 0.20 |
| Vehicle damage severity | | | | |
| Light | 47 (22.4%) | 43 (32.3%) | 35 (26.5%) | 0.01 |
| Moderate | 58 (27.6%) | 37 (27.8%) | 29 (22.0%) | |
| Severe | 67 (31.9%) | 24 (18.0%) | 39 (29.5%) | |
| Demolished | 19 (9.0%) | 7 (5.3%) | 8 (6.1%) | |
| None | 8 (3.8%) | 7 (5.3%) | 8 (6.1%) | |
| Other/unknown | 11 (5.2%) | 15 (11.3%) | 13 (9.8%) | |
| Crash year | | | | |
| 2010 | 44 (21.0%) | 22 (16.5%) | 20 (15.2%) | 0.33 |
| 2011 | 32 (15.2%) | 33 (24.8%) | 26 (19.7%) | |
| 2012 | 49 (23.3%) | 31 (23.3%) | 27 (20.5%) | |
| 2013 | 29 (13.8%) | 16 (12.0%) | 30 (22.7%) | |
| 2014 | 25 (11.9%) | 16 (12.0%) | 15 (11.4%) | |
| 2015 | 27 (12.9%) | 11 (8.3%) | 12 (9.1%) | |
| 2016 | < 5 | < 5 | < 5 | |
| Crash season | | | | |
| Winter (Dec-Feb) | 53 (25.2%) | 48 (36.1%) | 38 (28.8%) | 0.09 |
| Spring (Mar-May) | 52 (24.8%) | 31 (23.3%) | 27 (20.5%) | |
| Summer (Jun-Aug) | 54 (25.7%) | 22 (16.5%) | 39 (29.5%) | |
| Fall (Sep-Nov) | 51 (24.3%) | 32 (24.1%) | 28 (21.2%) | |
| Day of week | | | | |
| Weekday (Mon-Thur) | 119 (56.7%) | 75 (56.4%) | 91 (68.9%) | 1.00 |
| Weekend (Fri-Sun) | 91 (43.3%) | 58 (43.6%) | 41 (31.1%) | |
| Time of day | | | | |
| Morning (06:01-12:00) | 47 (22.4%) | 25 (18.8%) | 34 (25.8%) | 0.70 |
| Afternoon (12:01-18:00) | 107 (51.0%) | 70 (52.6%) | 63 (47.7%) | |
| Evening (18:01-21:00) | 32 (15.2%) | 17 (12.8%) | 14 (10.6%) | |
| Night (21:01-06:00) | 21 (10.0%) | 19 (14.3%) | 15 (11.4%) | |
| Other/unknown | < 5 | < 5 | 6 (4.5%) | |

**Legend:** Table depicting crash characteristics for responsible and non-responsible drivers. We had detailed police-reported crash data for all syncope cohort members but lacked crash data for other drivers involved in the crash. As a result, except where the index driver's data directly suggested contribution from others (e.g. pedestrian error, previous traffic crash), the 'contributions from other parties' factor could not account for other drivers' actions. This may have biased responsibility scores downward and effect estimates toward the null. However, the proportion of crash-involved drivers deemed responsible was similar to that within the largest prior responsibility study (44.2% versus 46%, respectively), suggesting missing data on 'contributions from other parties' had a limited effect on our results.[1]

Not all displayed data are components of the responsibility score tool.

As expected, responsible drivers were more likely to be disobeying road laws and were more likely to be involved in crashes that occurred on dry roads, during optimal weather, in full daylight, and involving only a single vehicle (these are components of the responsibility score).

Reassuringly, established risk factors for crash that are not part of the responsibility score are also more common among responsible drivers. For example, human contributory factors, which include alcohol, fatigue, and distraction/inattention, were more common among responsible than among non-responsible drivers (54.3% vs 16.5%; unadjusted odds ratio, 6.00; 95%CI, 3.51 to 10.2; p<0.001).

Other established risk factors for crash presented in Table 1 that are not a part of the responsibility score were also associated with crash responsibility, including male sex (62.9% of responsible drivers and 50.4% of non-responsible drivers; unadjusted odds ratio, 1.67; 95%CI, 1.07 to 2.59; p=0.025) and a prior history of any traffic contravention in a 5-year lookback (68.1% vs 51.9%; unadjusted odds ratio, 1.98; 95%CI, 1.27 to 3.10; p=0.003).

**Item S7: Forest plot of subgroup analyses**

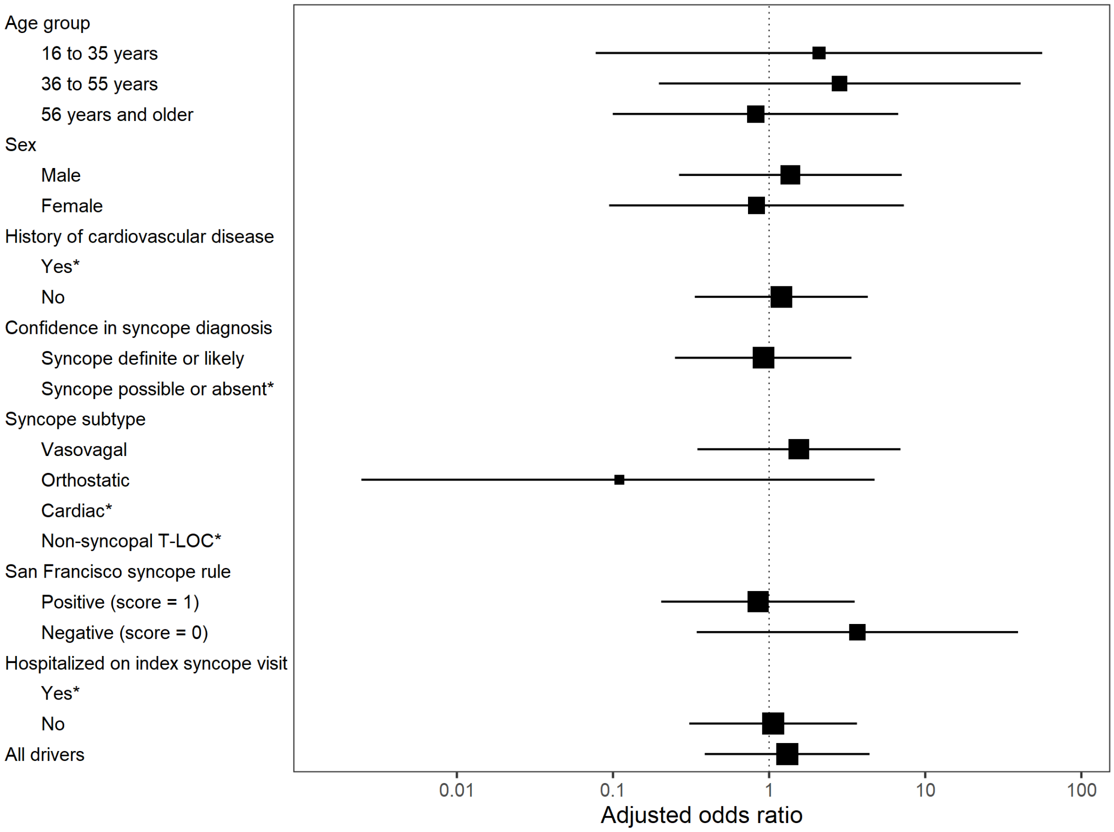


**Legend:** X-axis depicts the adjusted odds ratio for the association between syncope and crash responsibility; y-axis, the subgroup; square points, the adjusted odds ratio point estimate (with size reflecting the inverse of the standard error); horizontal lines, the 95% confidence interval, with arrow heads indicating the confidence interval endpoint is beyond the limit of the x-axis. Syncope was not associated with crash responsibility in any subgroup. *Indicates that all drivers in these strata were deemed responsible for their crash, making it impossible to calculate an odds ratio.
